# Increased risk of psychiatric sequelae of COVID-19 is highest early in the clinical course

**DOI:** 10.1101/2021.11.30.21267071

**Authors:** Ben Coleman, Elena Casiraghi, Hannah Blau, Lauren Chan, Melissa Haendel, Bryan Laraway, Tiffany J Callahan, Rachel R Deer, Ken Wilkins, Justin Reese, Peter N Robinson

## Abstract

**Background:** COVID-19 has been shown to increase the risk of adverse mental health consequences. A recent electronic health record (EHR)-based observational study showed an almost two-fold increased risk of new-onset mental illness in the first 90 days following a diagnosis of acute COVID-19.

**Methods:** We used the National COVID Cohort Collaborative, a harmonized EHR repository with 2,965,506 COVID-19 positive patients, and compared cohorts of COVID-19 patients with comparable controls. Patients were propensity score-matched to control for confounding factors. We estimated the hazard ratio (COVID-19:control) for new-onset of mental illness for the first year following diagnosis. We additionally estimated the change in risk for new-onset mental illness between the periods of 21-120 and 121-365 days following infection.

**Findings:** We find a significant increase in incidence of new-onset mental disorders in the period of 21-120 days following COVID-19 (3.8%, 3.6-4.0) compared to patients with respiratory tract infections (3%, 2.8-3.2). We further show that the risk for new-onset mental illness decreases over the first year following COVID-19 diagnosis compared to other respiratory tract infections and demonstrate a reduced (non-significant) hazard ratio over the period of 121-365 days following diagnosis. Similar findings are seen for new-onset anxiety disorders but not for mood disorders.

**Interpretation:** Patients who have recovered from COVID-19 are at an increased risk for developing new-onset mental illness, especially anxiety disorders. This risk is most prominent in the first 120 days following infection.

**Funding:** National Center for Advancing Translational Sciences (NCATS).

## INTRODUCTION

Severe acute respiratory syndrome coronavirus 2 (SARS-CoV-2), the virus that causes coronavirus disease 2019 (COVID-19), is responsible for nearly 5 million deaths worldwide and continues to cause over 200,000 deaths a month.^1^ COVID-19 is associated with a wide range of clinical manifestations that are still only partially understood.^2^ Recent reports have documented that a large proportion of COVID-19 patients develop neuropsychiatric symptoms during or after acute infection.^3^

Evidence suggests an increased risk of mental illness following viral infections,^4,5^ although definitive proof as well as a detailed understanding of molecular mechanisms are lacking. Viral infections resulting from recent outbreaks of Severe Acute Respiratory Syndrome (SARS) in 2002 and Middle East Respiratory Syndromes (MERS) in 2012, both caused by coronaviruses closely related to SARS-CoV-2, were associated with neurological manifestations in some cases.^6–9^ Several studies have also found evidence for increased risk of mental illness following SARS-CoV-2 infection.^3,10–19^

It is important to understand the risk of long-term psychiatric manifestations following COVID-19 because even a small increase in risk would have major public health ramifications. A study on a cohort of 44,779 COVID-19 patients and propensity score-matched controls from the TriNetX network found a statistically significant increase in new-onset mental illness 14 to 90 days following diagnosis for patients who had COVID-19 compared to one of the control health events.^11^ This finding was subsequently extended to a larger cohort of 236,379 COVID-19 patients and an observation period of up to 6 months with comparable results.^19^

To address these questions, it is critical to analyze a high volume of reliable patient-level, nationally representative data. In this study, we leveraged data from the National COVID Cohort Collaborative (N3C) (covid.cd2h.org), a centralized, harmonized, high-granularity electronic health record (EHR) repository.^20^ We confirmed the above-cited findings of increased risk for 46,610 COVID-19 survivors to develop a mental illness within 120 days following acute infection. We followed study participants up to 365 days, and assessed the difference in risk during the period of 121-365 days.

## METHODS

### Study Design

In this retrospective cohort study, we examine the incidence of new-onset mental illness for patients who have recovered from COVID-19 compared to control patients with a similar health event. We used patient data provided by the N3C accessed through the National Institute of Health (NIH) N3C Data Enclave. N3C has harmonized EHRs from 65 clinical organizations in the United States. 14 sites were removed because of missing BMI data. Records from the remaining 51 sites were frozen on October 20, 2021, and comprised data from 7,139,696 patients, of whom 1,834,913 were COVID-19 positive. Data were available for over 4.6 billion lab results, 1.4 billion drug exposures, and 469 million procedures from 467 million healthcare encounters. Data are collected from the clinical organizations, normalized to the Observational Medical Outcomes Partnership (OMOP) 5.3.1 vocabulary, then de-identified and made available to participating N3C research institutions. The study was exempted by the Institutional Review Board (IRB) at the Jackson Laboratory under 45 CFR 46.101(b) (Common Rule). The N3C data transfer to the National Center for Advancing Translational Sciences (NCATS) was approved by Johns Hopkins Medicine IRB-3 as single IRB of Protocol # IRB00249128 and individual site agreements with NIH.

### Study Variables and Outcomes

Clinical data including comorbidities, medications, and outcomes were identified using concept identifiers in the OMOP common data model. Demographics, laboratory values, COVID-19 status, and psychiatric diagnoses were collected for each patient.

Patients were included in the primary analysis if they had a confirmed diagnosis of SARS-CoV-2 by polymerase chain reaction (PCR) or antigen test after January 1, 2020. Patients with suspected COVID-19 who did not have a confirmatory test were excluded from this study. We defined three controls of comparable health events to COVID-19. Controls and outcomes were defined using codesets derived from OMOP standard vocabulary hierarchies, taking the diagnosis of interest and all known descendants in OMOP. Controls included respiratory tract infection (RTI; concept id: 4170143), large bone fractures (concept ids: 4129394, 4300192, 46270317, 4278672, 442619, 4205238, 442560, 4185758, 4059173), and urolithiasis (concept id: 4319447). Control patients were not restricted by the date of initial presentation. Patients with a history of any mental illness (concept ids: 432586, 441542) prior to 21 days after COVID-19 diagnosis and patients without a medical record extending back a year prior to COVID-19 were excluded from this analysis.

The date when a patient first came to medical attention for COVID-19 or for one of the control health conditions (RTI, fractures, or urolithiasis) will be referred to as the “initial presentation” in the following sections. Psychiatric outcomes included any mental disorder (concept ids: 432586, 441542), mood disorder (concept id: 444100), and anxiety disorder (concept id: 441542). Dyspnea (concept id: 312437) was included as a positive control.

#### Analysis of Psychiatric Sequelae

Data analysis was performed using Palantir Foundry (Palantir Technologies Inc., Denver, Colorado). The analysis was structured as a directed acyclic graph of data transformations by leveraging the capabilities of the Palantir platform. Individual transformations were implemented as nodes consisting of SQL, Python, or R code.

Patients were matched with patients from their reporting institution within 5 years of age. Propensity score matching was done on 34 factors using a logistic regression model including main effect terms. The factors comprised age, race, ethnicity, gender, smoking status, Charlson Comorbidity Score, BMI, visit type, length of stay (if applicable), cerebral infarction, chronic respiratory disease, type 1 diabetes, type 2 diabetes, ischemic heart disease, nonischemic heart disease, chronic hepatitis, alcoholic liver damage, hepatic failure, hepatic fibrosis, hepatic passive congestion, hepatic steatosis, portal hypertension, other liver disease, hypertension, hypertensive kidney disease, nonhypertensive chronic kidney disease, lymphoid neoplasm, other neoplasm, lupus, rheumatoid arthritis, rheumatoid arthritis with rheumatoid factor, psoriasis, nicotine dependence, and immunosuppression. All comorbidities were defined as a binary variable indicating whether that patient had received the diagnosis of the comorbidity prior to initial presentation. Patients with missing data on any of the covariates were excluded from the analysis. BMI was the most commonly missing variable (57.6%) followed by age (0.98%) and length of stay (0.043%). Little’s test^21^ was used to characterize patterns of missingness, and results brought no evidence of data being missing completely at random. Propensity score matching with a greedy nearest neighbor approach was used to match patients. Patients were matched at a ratio of 1:1 for RTI and fracture and 2:1 for the urolithiasis control group since there were fewer patients with urolithiasis. Any predictor with a standardized mean difference between cohorts lower than 0.1 was considered well matched (S3).^18^

Multivariable Cox regression was performed to compare the incidence of new-onset mental illness for all psychiatric conditions, mood disorders, and anxiety disorders for 21 to 365 days following initial presentation. For each patient, the latest visit and date of death (if observed) were recorded to measure the time to right censoring (the last date for which the outcome can be ruled out) and survival, respectively. Time to event (diagnosis of a psychiatric condition) was defined with respect to the initial presentation as described above. The *survival* R package^22^ was used to produce Kaplan-Meier estimated survival curves, to compute Cox proportional hazards regression, and to compute and visualize Schoenfeld residuals.

A secondary analysis was performed to assess a potential relevance of the number of visits following COVID or RTI. The number of visits was counted up to the final event in the survival curve (either diagnosis of a mental illness or the last day of observation), and was divided the the total number of days between day 21 and the final event.

#### Analysis of time-varying hazard ratio

We tested the proportional hazard assumption for the Cox regression for comparisons of COVID-19 and controls.^23^ An apparent inflection point of the survival curve was observed empirically on the plot of the log-log cumulative hazard for each group. Cox regression-based estimation of the hazard ratio was therefore conducted over two separate intervals, days 21-120 and 121-365.

#### Code availability

The analysis was implemented using SQL, Python, and R and is documented in the Supplemental material.

## Results

A total of 7,139,696 patients in the N3C Data Enclave were assessed in our data set frozen on October 20, 2021. Of these patients, 1,834,913 had a positive COVID-19 PCR or antigen test. We restricted analysis to patients with no previous recorded psychiatric illness and at least one year of data prior to acute COVID-19 (or control health event). This left 638,121 patients in the COVID-19 cohort. Control cohorts were formed based on the following health events: RTI other than COVID-19, large bone fracture, and urolithiasis. We focused our analysis on RTI as this control group had the most patients and is the most comparable to COVID-19. The median age of all groups combined was 42 years and 411,640 were female (Table 1).

**Table 1.**
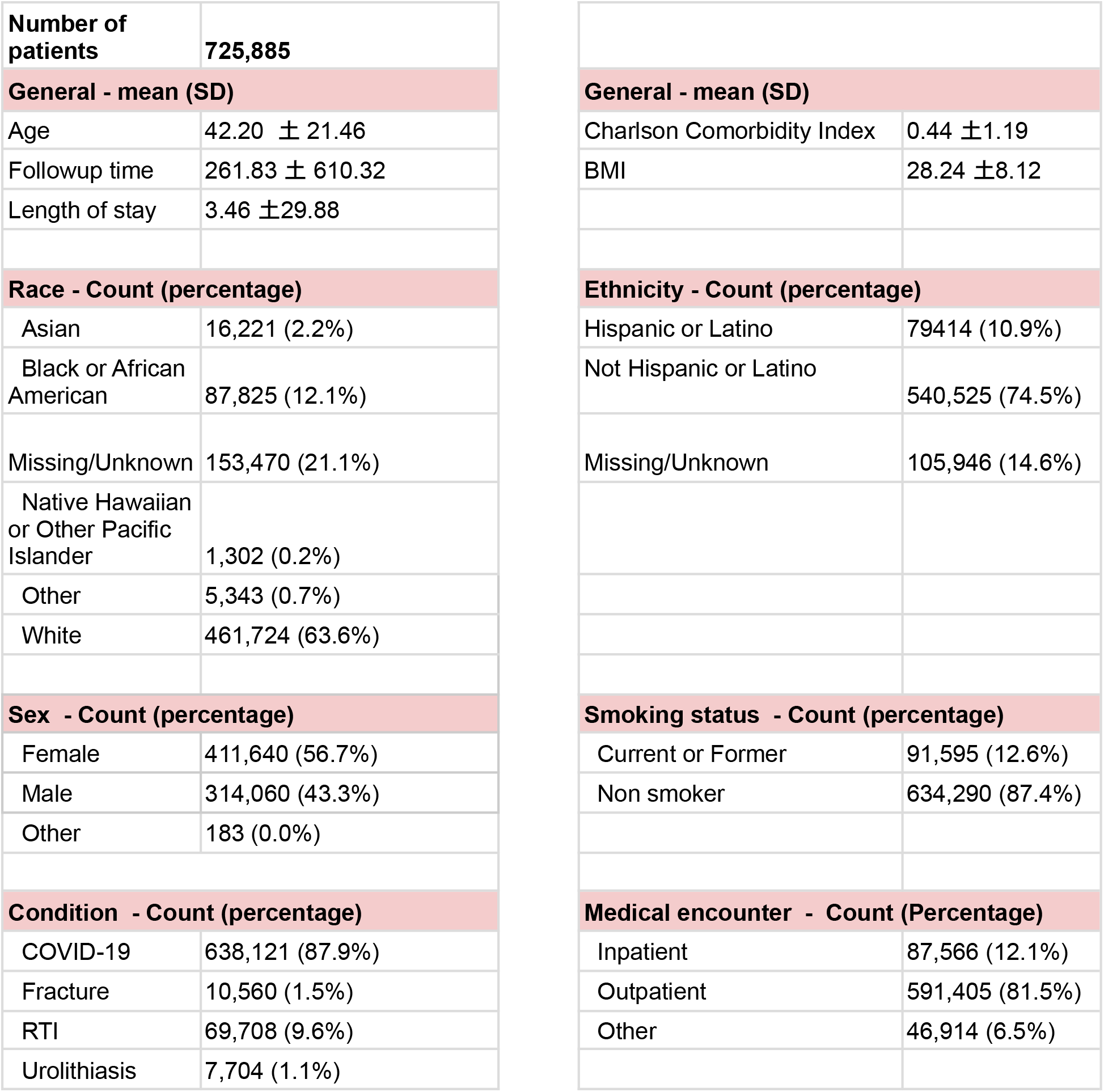
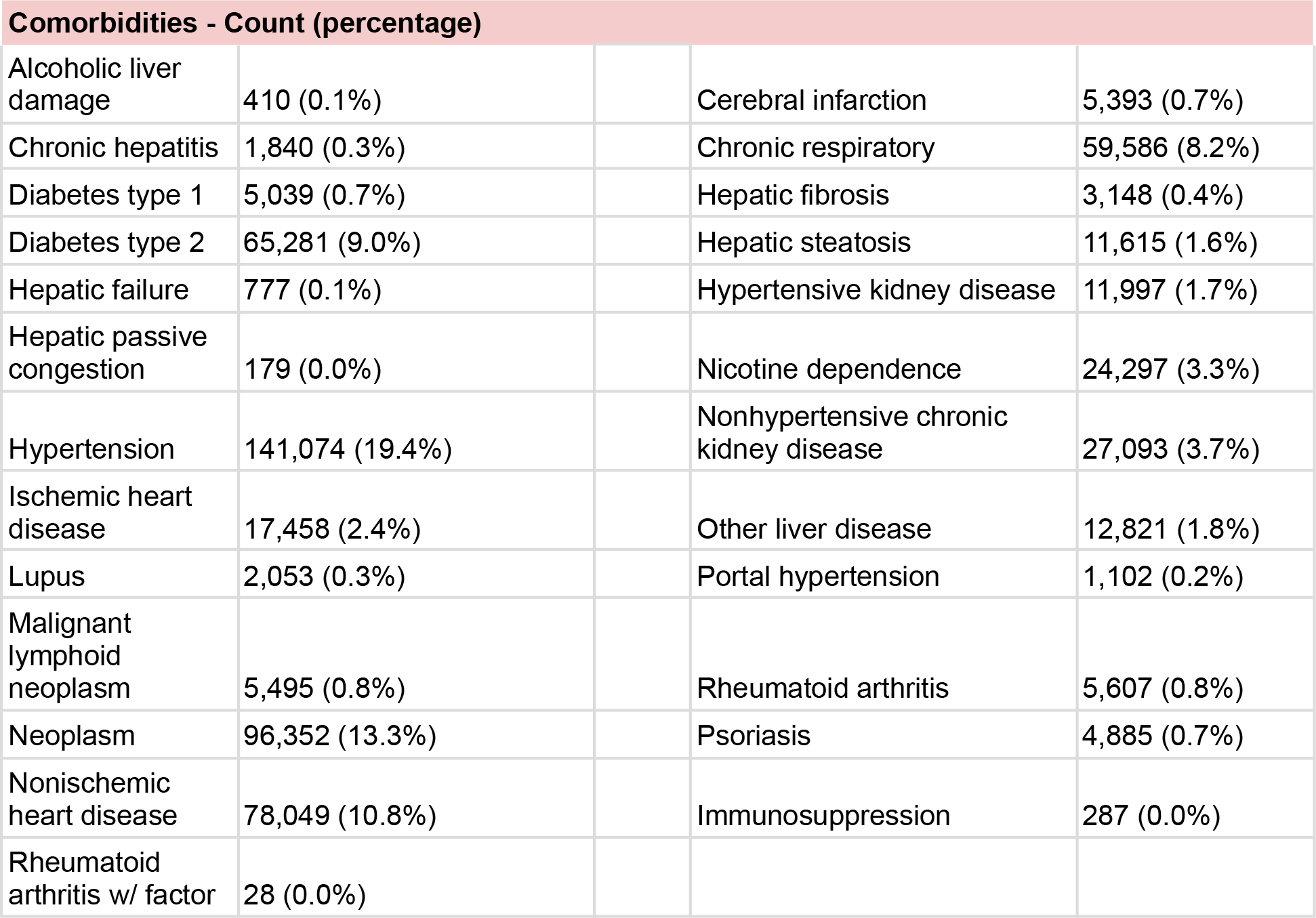
Characteristics of the study cohort are presented as counts (with percentage for categorical/boolean variables) or as mean ± SD (for numeric variables). The percentage is calculated with respect to the size of the entire group (725,885), which is all patients with one or more of COVID-19, fracture, RTI (respiratory tract infection other than COVID-19), or urolithiasis.

|  |  |
| --- | --- |
| **Number of patients** | **725,885** |
| **General - mean (SD)** |  |
| Age | 42.20 ± 21.46 |
| Followup time | 261.83 ± 610.32 |
| Length of stay | 3.46 ± 29.88 |
| **Race - Count (percentage)** |  |
| Asian | 16,221 (2.2%) |
| Black or African American | 87,825 (12.1%) |
| Missing/Unknown | 153,470 (21.1%) |
| Native Hawaiian or Other Pacific Islander | 1,302 (0.2%) |
| Other | 5,343 (0.7%) |
| White | 461,724 (63.6%) |
| **Sex - Count (percentage)** |  |
| Female | 411,640 (56.7%) |
| Male | 314,060 (43.3%) |
| Other | 183 (0.0%) |
| **Condition - Count (percentage)** |  |
| COVID-19 | 638,121 (87.9%) |
| Fracture | 10,560 (1.5%) |
| RTI | 69,708 (9.6%) |
| Urolithiasis | 7,704 (1.1%) |
| **General - mean (SD)** |  |
| Charlson Comorbidity Index | 0.44 ± 1.19 |
| BMI | 28.24 ± 8.12 |
| **Ethnicity - Count (percentage)** |  |
| Hispanic or Latino | 79414 (10.9%) |
| Not Hispanic or Latino | 540,525 (74.5%) |
| Missing/Unknown | 105,946 (14.6%) |
| **Smoking status - Count (percentage)** |  |
| Current or Former | 91,595 (12.6%) |
| Non smoker | 634,290 (87.4%) |
| **Medical encounter - Count (Percentage)** |  |
| Inpatient | 87,566 (12.1%) |
| Outpatient | 591,405 (81.5%) |
| Other | 46,914 (6.5%) |

| Comorbidities - Count (percentage) |  |  |  |
| --- | --- | --- | --- |
| Alcoholic liver damage | 410 (0.1%) | Cerebral infarction | 5,393 (0.7%) |
| Chronic hepatitis | 1,840 (0.3%) | Chronic respiratory | 59,586 (8.2%) |
| Diabetes type 1 | 5,039 (0.7%) | Hepatic fibrosis | 3,148 (0.4%) |
| Diabetes type 2 | 65,281 (9.0%) | Hepatic steatosis | 11,615 (1.6%) |
| Hepatic failure | 777 (0.1%) | Hypertensive kidney disease | 11,997 (1.7%) |
| Hepatic passive congestion | 179 (0.0%) | Nicotine dependence | 24,297 (3.3%) |
| Hypertension | 141,074 (19.4%) | Nonhypertensive chronic kidney disease | 27,093 (3.7%) |
| Ischemic heart disease | 17,458 (2.4%) | Other liver disease | 12,821 (1.8%) |
| Lupus | 2,053 (0.3%) | Portal hypertension | 1,102 (0.2%) |
| Malignant lymphoid neoplasm | 5,495 (0.8%) | Rheumatoid arthritis | 5,607 (0.8%) |
| Neoplasm | 96,352 (13.3%) | Psoriasis | 4,885 (0.7%) |
| Nonischemic heart disease | 78,049 (10.8%) | Immunosuppression | 287 (0.0%) |
| Rheumatoid arthritis w/ factor | 28 (0.0%) |  |  |

The outcomes of interest were the psychiatric conditions: any mental disorder, mood disorder, and anxiety disorder. The outcome of dyspnea, whose prevalence is expected to be increased following COVID-19,^15^ was used as positive controls.

These conditions were considered sequelae of COVID-19 if they occurred at least 21 days after the diagnosis of COVID-19. We fitted a sequence of Cox regression models to estimate and assess the significance of the association between patients having a psychiatric sequela and belonging to the COVID-19 group compared to one of the control groups. This gave us a hazard ratio estimate for each comparison based on the respective propensity-matched cohorts. We also used the fitted Cox regression model to test the proportional hazard assumption^23^ of each included variable.

Schoenfeld residual analysis yielded a significant p-value and led us to reject the null hypothesis of a constant proportional hazard over the full time period of 21-365 days for our primary control group (RTI). We therefore separated the cohort into two time intervals (before and after 120 days) in which the proportional hazard assumption was not violated. Table 2 provides the estimated probabilities of experiencing a psychiatric sequela of COVID-19 in days 21-120 or 21-365 following acute infection. We identified a statistically significant difference in the hazard rate of sequelae between COVID-19 and RTI in the early post-acute phase (from 21 to 120 days) but not in the late post-acute phase (from 121 to 365 days). The estimated probability (as modeled on the log-hazard scale over time) of a new-onset psychiatric diagnosis in the early post-acute phase was significantly higher for the COVID-19 group than for the RTI group (HR 1.3, 1.2-1.4). The hazard ratio for new-onset mental illness in the late phase was not significantly different in COVID-19 compared to the RTI group.

**Table 2.** Estimated incidences of new-onset mental illness are shown for COVID-19 and RTI (fracture and urolithiasis, Supplemental Tables S1 and S2) at 120 and 365 days following diagnosis. Hazard ratios, Wald-type Cox regression coefficient, and Grambsch-Therneau proportional hazard assumption tests are shown for the period of 21-120 days and 121-365 days. Dyspnea is shown as a positive control whose incidence is expected to be higher following COVID-19.

|  | COVID-19 | RTI | Hazard Ratio | Hazard Ratio P-Value | Proportional hazard p value |
| --- | --- | --- | --- | --- | --- |
| **21-120 Days (early post-acute phase)** |  |  |  |  |  |
| **All psychiatric illness** | 3.8%<br>(3.6-4.0) | 3.0%<br>(2.8-3.2) | 1.3 (1.2-1.4) | <0.0001 | 0.94 |
| **Mood disorder** | 1.2%<br>(1.1-1.3) | 1.1%<br>(1-1.2) | 1.1 (0.99-1.3) | 0.066 | 0.72 |
| **Anxiety** | 2.0%<br>(1.8-2.1) | 1.6%<br>(1.5-1.7) | 1.3 (1.1-1.4) | <0.0001 | 0.72 |
| **Dyspnea** | 3.7%<br>(3.5-3.9) | 2.8%<br>(2.7-3) | 1.4 (1.2-1.5) | <0.0001 | 0.38 |
| **121-365 Days (late post-acute phase)** |  |  |  |  |  |
| **All psychiatric illness** | 11%<br>(10.4-11.2) | 10%<br>(9.7-10.4) | 1 (0.97-1.1) | 0.29 | 0.049 |
| **Mood disorder** | 4% (3.7-4.3) | 3.7%<br>(3.5-3.9) | 1.1 (0.97-1.2) | 0.16 | 0.28 |
| **Anxiety** | 6% (5.7-6.3) | 5.7%<br>(5.4-5.9) | 1 (0.91-1.1) | 0.99 | 0.33 |
| **Dyspnea** | 7.8%<br>(7.5-8.2) | 6.5%<br>(6.2-6.8) | 1.2 (1-1.3) | 0.0039 | 0.10 |

Similar findings were obtained for anxiety and mood disorders. The estimated probability of a new-onset anxiety diagnosis was significantly increased for COVID-19 patients compared to RTI patients in the early post-acute phase (HR 1.3, 1.1-1.4). However, the estimated probability of a new-onset mood disorder diagnosis in the same period was not significantly increased for COVID-19 patients in comparison to RTI patients. Neither anxiety nor mood disorders were significantly increased in the interval of 121-365 days following initial presentation. In contrast, there was a significantly increased hazard ratio for dyspnea, a known post-acute COVID-19 sequela,^15^ in both time periods (Figure 1, Table 2).

**Figure 1:**
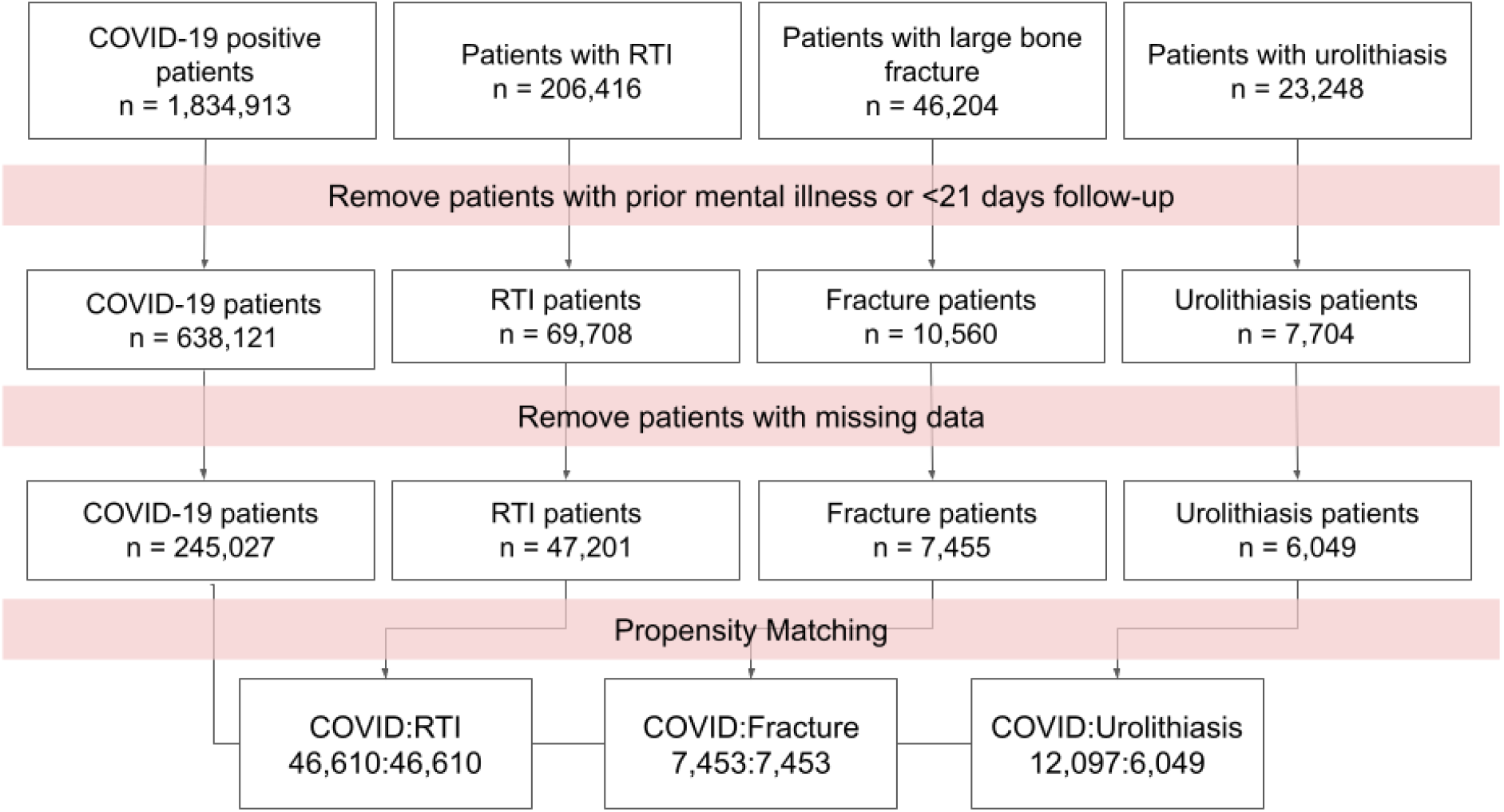
Workflow for creating matched patient groups within N3C. Four groups are created, patients with a positive PCR or antigen test for COVID-19 and patients with a diagnosis of one of the control events. Patients with mental illness prior to the post-COVID phase (21 days after diagnosis) are removed. Patients with missing data are removed. Finally, each control group is propensity matched with a group of COVID-19 patients in 1:1 or 2:1 ratios. RTI: Respiratory tract infection other than COVID-19.

**Figure 2.**
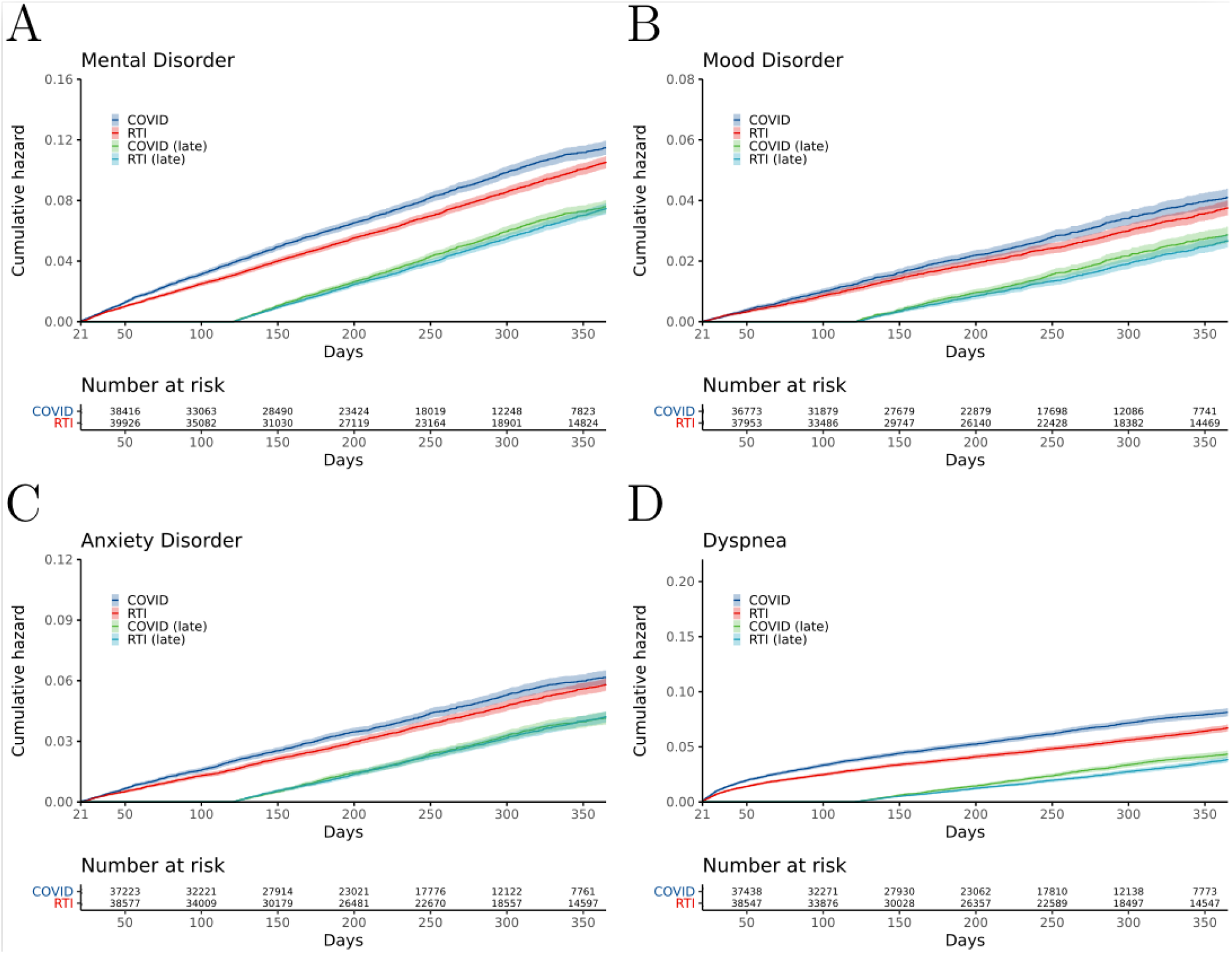
Cumulative incidence plots for new-onset psychiatric illness following acute COVID-19 compared to other respiratory tract infections (RTIs). Shaded areas represent 95% CIs. Number at risk refers to the number of subjects in each cohort who did not have the indicated outcome before the indicated number of days following the onset of COVID-19 or RTI. COVID (late) and RTI (late) refer to patients who did not receive a diagnosis in the first 120 days following initial presentation. A) All Mental Illness; B) Mood disorders; C) Anxiety disorder; D) Dyspnea (positive control). Similar cumulative incidence curves for additional events and outcomes are shown in the Supplemental Figures S1-S2.

Hospitalization for pneumonia is associated with an increased risk for depressive symptoms that can be observed even in persons without substantial medical comorbidities or with non-critical pneumonia.^24^ This observation motivated our choice of RTI as a control group. Similar observations have been made for urolithiasis and large-bone fracture.^25,26^ We therefore compared COVID-19 to these two cohorts and found that the hazard ratio was significantly increased in COVID-19 compared to these controls in both the early and late post-acute phases (HR 1.4 and between 1.2-1.5 respectively). (See Supplemental Tables S1-S2 and Supplemental Figures S2-S3).

We reasoned that patients might be followed more closely following COVID-19 as compared with other RTIs and that a higher visit frequency might increase the probability of a mental illness being recorded in the medical records. To assess this, we repeated our analysis but added the frequency of visits 21 days or more after initial presentation as a factor to the Cox regression. We found that the HR of getting any mental illness in the early post-acute phase was still significant (p value <0.0001), but reduced to 1.2 (1.1-1.3; Supplemental Table S3).

## Discussion

In this retrospective observational study on a cohort of 638,121 individuals following acute COVID-19 infection, we identified a statistically significant association of COVID-19 with the occurrence of new-onset psychiatric conditions. Our results confirm those of a previously published study on 44,779 COVID-19 patients in the TriNetX network,^11^ but the degree of increased risk was less. For instance, the point estimate for new-onset psychiatric diagnoses during the first 14 to 90 days was 5.8% after a diagnosis of COVID-19 as compared to 3.4% after a diagnosis of another RTI in the previous study.^11^ In our study, the point estimate for new-onset psychiatric diagnoses during the first 21 to 120 days was 3.8% after a diagnosis of COVID-19 as compared to 3.0% after a diagnosis of another RTI. Our analysis did not find a significantly increased risk of new-onset psychiatric diagnoses in the period from 121 to 365 days when compared to RTIs. However, we did identify an association of COVID-19 with increased risk across the entire time period when compared to urolithiasis and fracture, suggesting there might be a spectrum of increased risk for mental illness following different diseases.

There are several potential reasons for the differences between our results and those of the above-cited study. The previous study included data from the beginning of the pandemic on January 20, 2020 (date of the first recorded COVID-19 case in the USA) to August 1, 2020, while our study includes data up to October 20, 2021. It is conceivable that perceptions of COVID-19 by patients could have shifted in the intervening time or that clinical practice has changed in the subsequent year. It is possible that improved treatment options available later in the pandemic have reduced the increased risk of psychiatric illness. Finally, it has been reported that COVID-19 vaccination may reduce rates of anxiety and depression and alleviate symptoms in persons with long COVID,^27,28^ and thus it is possible that the increasing availability of vaccines has reduced the rate of mental illness following COVID-19. The data available in N3C does not include comprehensive information about vaccination status, so we cannot test this hypothesis. Our study has several strengths, including the large sample size and relatively long follow-up time. Our dataset is derived from over 50 institutions across the country with over 1.8 million cases of COVID-19, and thus is a representative sample of the COVID-19 positive population in the United States. Limitations of our study include the limited amount of information available prior to the acute COVID-19 event (median of 978 days), meaning that some portion of the events we classified as new-onset could be recurrences of a prior mental illness recorded before the period for which data were available in N3C. Samples with missing data were removed, which reduced the size of the cohort used for analysis. In addition to missingness, N3C data may be influenced by the heterogeneity of coding practices at the participating institutions related to the use of four different federated common data models and local coding practices. N3C employs a comprehensive suite of data quality checks to mitigate this issue,^29^ but residual problems cannot be ruled out. Our decision to perform Cox regression analysis separately on the early and late periods was made post hoc, after we determined that the assumption of proportional hazard did not hold for the entire time period. That said, anyone subsequently comparing our resulting estimates to those reported by other studies ought to bear in mind the apples-and-oranges issue inherent to Cox regression as conventionally used across studies: the non-collapsibility of the hazard ratio as a measure of exposure-outcome association, namely that estimates target distinctly different quantities when Cox models include distinct sets of covariates.^30^

## Conclusion

Our study supports previously published reports of an increased risk of new-onset psychiatric illness following acute COVID-19. In contrast to the nearly doubled risk identified by the previous study, we found the relative risk to be increased by only about 25% (3.8% vs. 3.0% following other RTI). We did not find a significant difference in risk in the time period of 121-365 days following acute COVID-19, suggesting that the increased risk of psychiatric illness is concentrated relatively early in the post-acute course. Our results have important implications for understanding the natural history of neuropsychiatric manifestations of COVID-19. If confirmed by independent studies, our findings suggest that to cope with the excess in psychiatric morbidity experienced by survivors of COVID-19, health services should focus efforts early in the post-COVID clinical course.

## Data Availability

The data presented in this paper can be accessed upon application to the NCATS N3C Data Enclave covid.cd2h.org/enclave.

## Data sharing

The data presented in this paper can be accessed upon application to the NCATS N3C Data Enclave at https://covid.cd2h.org/enclave.

## Acknowledgments

The analyses described in this publication were conducted with data or tools accessed through the NCATS N3C Data Enclave covid.cd2h.org/enclave and supported by NCATS U24 TR002306. This research was possible because of the patients whose information is included within the data from participating organizations (covid.cd2h.org/dtas) and the organizations and scientists (covid.cd2h.org/duas) who have contributed to the on-going development of this community resource (cite this https://doi.org/10.1093/jamia/ocaa196).

The N3C data transfer to NCATS is performed under a Johns Hopkins University Reliance Protocol # IRB00249128 or individual site agreements with NIH. The N3C Data Enclave is managed under the authority of the NIH; information can be found at https://ncats.nih.gov/n3c/resources.

We gratefully acknowledge contributions from the following N3C core teams:

(Asterisks indicate leads) • Principal Investigators: Melissa A. Haendel^*^, Christopher G. Chute^*^, Kenneth R. Gersing, Anita Walden

- Workstream, subgroup and administrative leaders: Melissa A. Haendel^*^, Tellen D. Bennett, Christopher G. Chute, David A. Eichmann, Justin Guinney, Warren A. Kibbe, Hongfang Liu, Philip R.O. Payne, Emily R. Pfaff, Peter N. Robinson, Joel H. Saltz, Heidi Spratt, Justin Starren, Christine Suver, Adam B. Wilcox, Andrew E. Williams, Chunlei Wu
- Key liaisons at data partner sites
- Regulatory staff at data partner sites
- Individuals at the sites who are responsible for creating the datasets and submitting data to N3C • Data Ingest and Harmonization Team: Christopher G. Chute^*^, Emily R. Pfaff^*^, Davera Gabriel, Stephanie S. Hong, Kristin Kostka, Harold P. Lehmann, Richard A. Moffitt, Michele Morris, Matvey B. Palchuk, Xiaohan Tanner Zhang, Richard L. Zhu
- Phenotype Team (Individuals who create the scripts that the sites use to submit their data, based on the COVID and Long COVID definitions): Emily R. Pfaff^*^, Benjamin Amor, Mark M. Bissell, Marshall Clark, Andrew T. Girvin, Stephanie S. Hong, Kristin Kostka, Adam M. Lee, Robert T. Miller, Michele Morris, Matvey B. Palchuk, Kellie M. Walters
- Project Management and Operations Team: Anita Walden^*^, Yooree Chae, Connor Cook, Alexandra Dest, Racquel R. Dietz, Thomas Dillon, Patricia A. Francis, Rafael Fuentes, Alexis Graves, Julie A. McMurry, Andrew J. Neumann, Shawn T. O’Neil, Usman Sheikh, Andréa M. Volz, Elizabeth Zampino
- Partners from NIH and other federal agencies: Christopher P. Austin^*^, Kenneth R. Gersing^*^, Samuel Bozzette, Mariam Deacy, Nicole Garbarini, Michael G. Kurilla, Sam G. Michael, Joni L. Rutter, Meredith Temple-O’Connor
- Analytics Team (Individuals who build the Enclave infrastructure, help create codesets, variables, and help Domain Teams and project teams with their datasets): Benjamin Amor^*^, Mark M. Bissell, Katie Rebecca Bradwell, Andrew T. Girvin, Amin Manna, Nabeel Qureshi
- Publication Committee Management Team: Mary Morrison Saltz^*^, Christine Suver^*^, Christopher G. Chute, Melissa A. Haendel, Julie A. McMurry, Andréa M. Volz, Anita Walden
- Publication Committee Review Team: Carolyn Bramante, Jeremy Richard Harper, Wenndy Hernandez, Farrukh M Koraishy, Federico Mariona, Amit Saha, Satyanarayana Vedula

Stony Brook University — U24TR002306 • University of Oklahoma Health Sciences Center — U54GM104938: Oklahoma Clinical and Translational Science Institute (OCTSI) • West Virginia University U54GM104942: West Virginia Clinical and Translational Science Institute (WVCTSI) • University of Mississippi Medical Center — U54GM115428: Mississippi Center for Clinical and Translational Research (CCTR) • University of Nebraska Medical Center — U54GM115458: Great Plains IDeA-Clinical & Translational Research • Maine Medical Center — U54GM115516: Northern New England Clinical & Translational Research (NNE-CTR) Network • Wake Forest University Health Sciences — UL1TR001420: Wake Forest Clinical and Translational Science Institute • Northwestern University at Chicago — UL1TR001422: Northwestern University Clinical and Translational Science Institute (NUCATS) • University of Cincinnati — UL1TR001425: Center for Clinical and Translational Science and Training • The University of Texas Medical Branch at Galveston — UL1TR001439: The Institute for Translational Sciences • Medical University of South Carolina — UL1TR001450: South Carolina Clinical & Translational Research Institute (SCTR) • University of Massachusetts Medical School Worcester — UL1TR001453: The UMass Center for Clinical and Translational Science (UMCCTS) • University of Southern California UL1TR001855: The Southern California Clinical and Translational Science Institute (SC CTSI) • Columbia University Irving Medical Center — UL1TR001873: Irving Institute for Clinical and Translational Research • George Washington Children’s Research Institute — UL1TR001876: Clinical and Translational Science Institute at Children’s National (CTSA-CN) • University of Kentucky — UL1TR001998: UK Center for Clinical and Translational Science • University of Rochester — UL1TR002001: UR Clinical & Translational Science Institute • University of Illinois at Chicago — UL1TR002003: UIC Center for Clinical and Translational Science • Penn State Health Milton S. Hershey Medical Center — UL1TR002014: Penn State Clinical and Translational Science Institute • The University of Michigan at Ann Arbor — UL1TR002240: Michigan Institute for Clinical and Health Research • Vanderbilt University Medical Center — UL1TR002243: Vanderbilt Institute for Clinical and Translational Research • University of Washington — UL1TR002319: Institute of Translational Health Sciences • Washington University in St. Louis — UL1TR002345: Institute of Clinical and Translational Sciences • Oregon Health & Science University — UL1TR002369: Oregon Clinical and Translational Research Institute • University of Wisconsin-Madison — UL1TR002373: UW Institute for Clinical and Translational Research • Rush University Medical Center — UL1TR002389: The Institute for Translational Medicine (ITM) • The University of Chicago — UL1TR002389: The Institute for Translational Medicine (ITM) • University of North Carolina at Chapel Hill — UL1TR002489: North Carolina Translational and Clinical Science Institute • University of Minnesota — UL1TR002494: Clinical and Translational Science Institute • Children’s Hospital Colorado — UL1TR002535: Colorado Clinical and Translational Sciences Institute • The University of Iowa — UL1TR002537: Institute for Clinical and Translational Science • The University of Utah — UL1TR002538: Uhealth Center for Clinical and Translational Science • Tufts Medical Center — UL1TR002544: Tufts Clinical and Translational Science Institute • Duke University — UL1TR002553: Duke Clinical and Translational Science Institute • Virginia Commonwealth University — UL1TR002649: C. Kenneth and Dianne Wright Center for Clinical and Translational Research • The Ohio State University UL1TR002733: Center for Clinical and Translational Science • The University of Miami Leonard M. Miller School of Medicine — UL1TR002736: University of Miami Clinical and Translational Science Institute • University of Virginia — UL1TR003015: iTHRIV Integrated Translational health Research Institute of Virginia • Carilion Clinic — UL1TR003015: iTHRIV Integrated Translational health Research Institute of Virginia • University of Alabama at Birmingham — UL1TR003096: Center for Clinical and Translational Science • Johns Hopkins University — UL1TR003098: Johns Hopkins Institute for Clinical and Translational Research • University of Arkansas for Medical Sciences — UL1TR003107: UAMS Translational Research Institute • Nemours — U54GM104941: Delaware CTR ACCEL Program • University Medical Center New Orleans — U54GM104940: Louisiana Clinical and Translational Science (LA CaTS) Center • University of Colorado Denver, Anschutz Medical Campus — UL1TR002535: Colorado Clinical and Translational Sciences Institute • Mayo Clinic Rochester — UL1TR002377: Mayo Clinic Center for Clinical and Translational Science (CCaTS) • Tulane University — UL1TR003096: Center for Clinical and Translational Science • Loyola University Medical Center — UL1TR002389: The Institute for Translational Medicine (ITM) • Advocate Health Care Network — UL1TR002389: The Institute for Translational Medicine (ITM) • OCHIN — INV-018455: Bill and Melinda Gates Foundation grant to Sage Bionetworks.

## Author contributions

Conceived and designed the analysis: BC, EC, KW, JR, PNR; implemented analysis, developed scripts: BC, JR, BL, HB, EC; Performed the analysis: BC, JR; Wrote the paper: BC, PNR. Critical revision: LC, MH, RD. All authors approved the final version.

Authorship was determined using ICMJE recommendations.

