## Supplement for "Increased risk of psychiatric sequelae of COVID-19 is highest early in the clinical course"

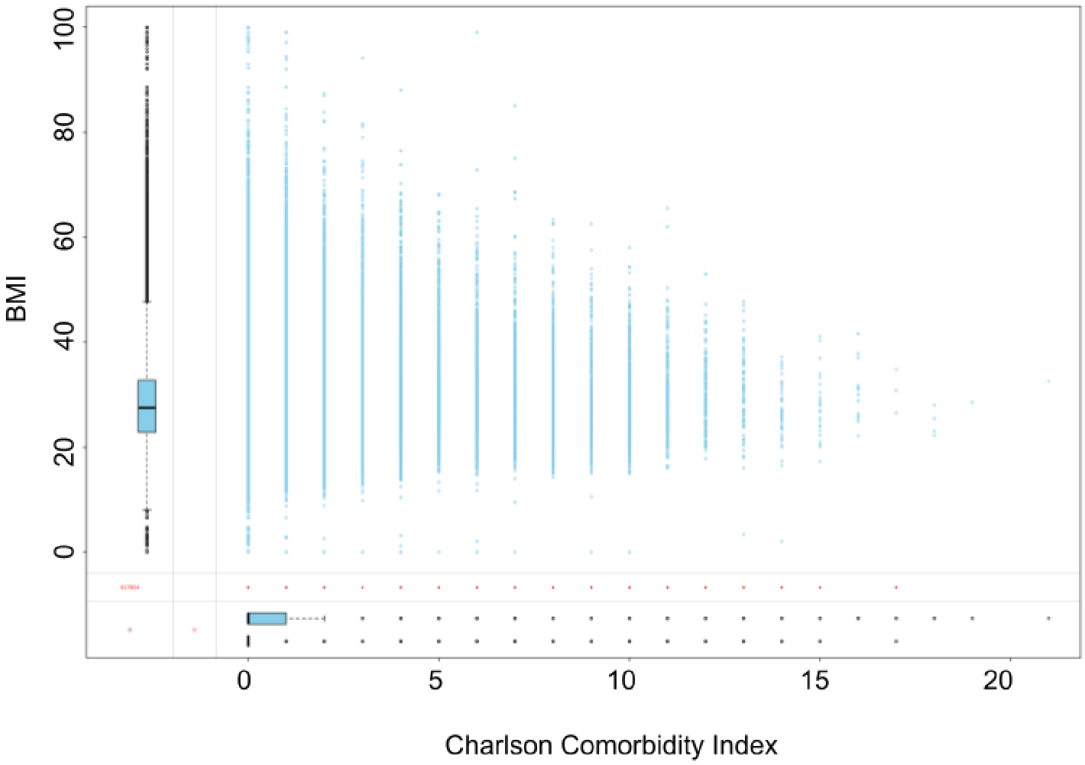


**Figure S1. Missingness analysis of BMI with respect to Charleson Comorbidity Index.** A box plot summarizing Charleson Comorbidity Index [1] of patients for whom BMI is missing (bottom box plot) or present (top box plot) is shown on the x-axis. A box plot summarizing BMI is shown on the y-axis. Graphic generated with the R VIM package [2]. There is no missing data for the Charlson Comorbidity Index. The plot suggests BMI data is missing completely at random.

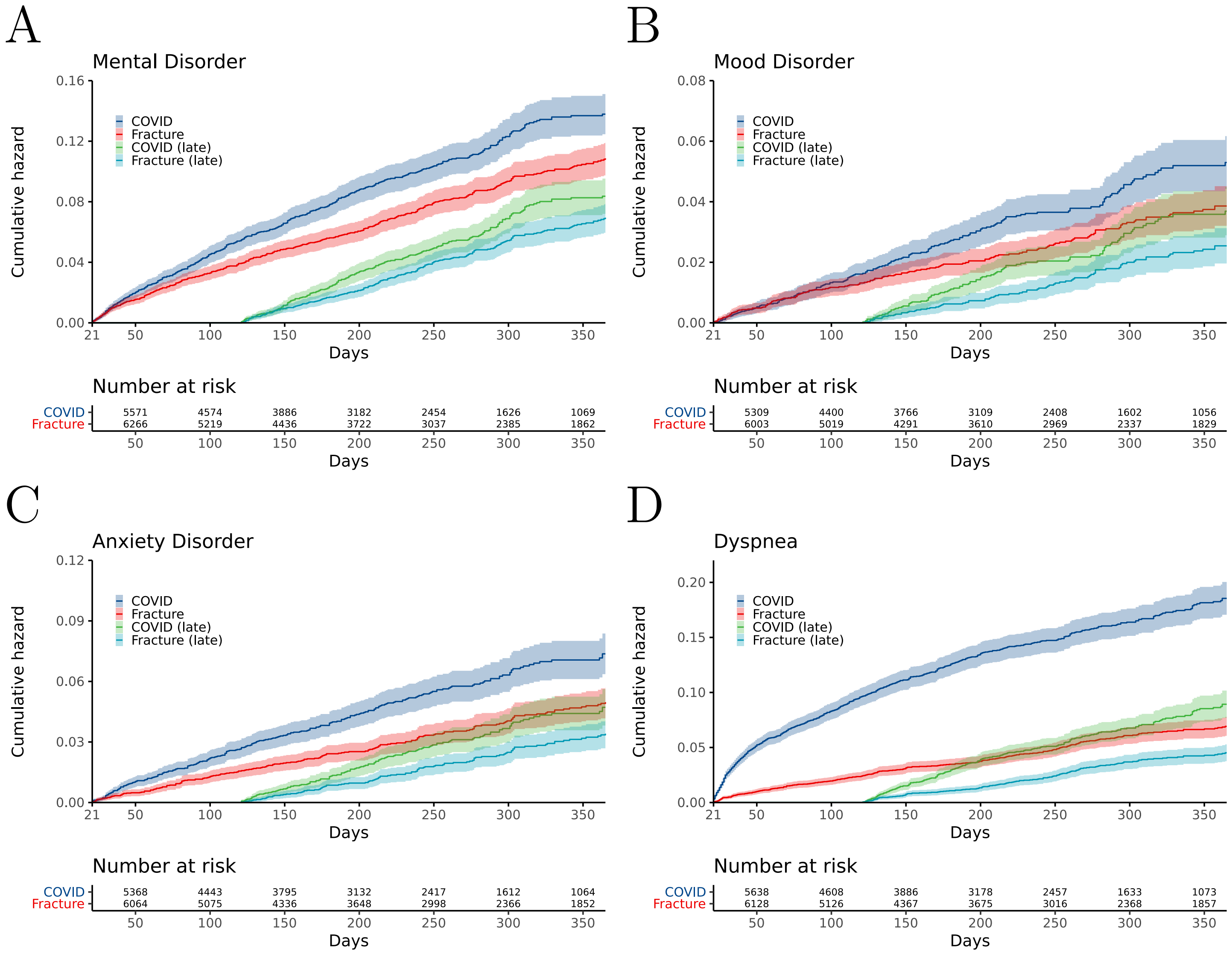


**Figure S2.** Kaplan-Meier curves for onset of first psychiatric diagnoses after COVID-19 diagnosis compared with large-bone fracture.

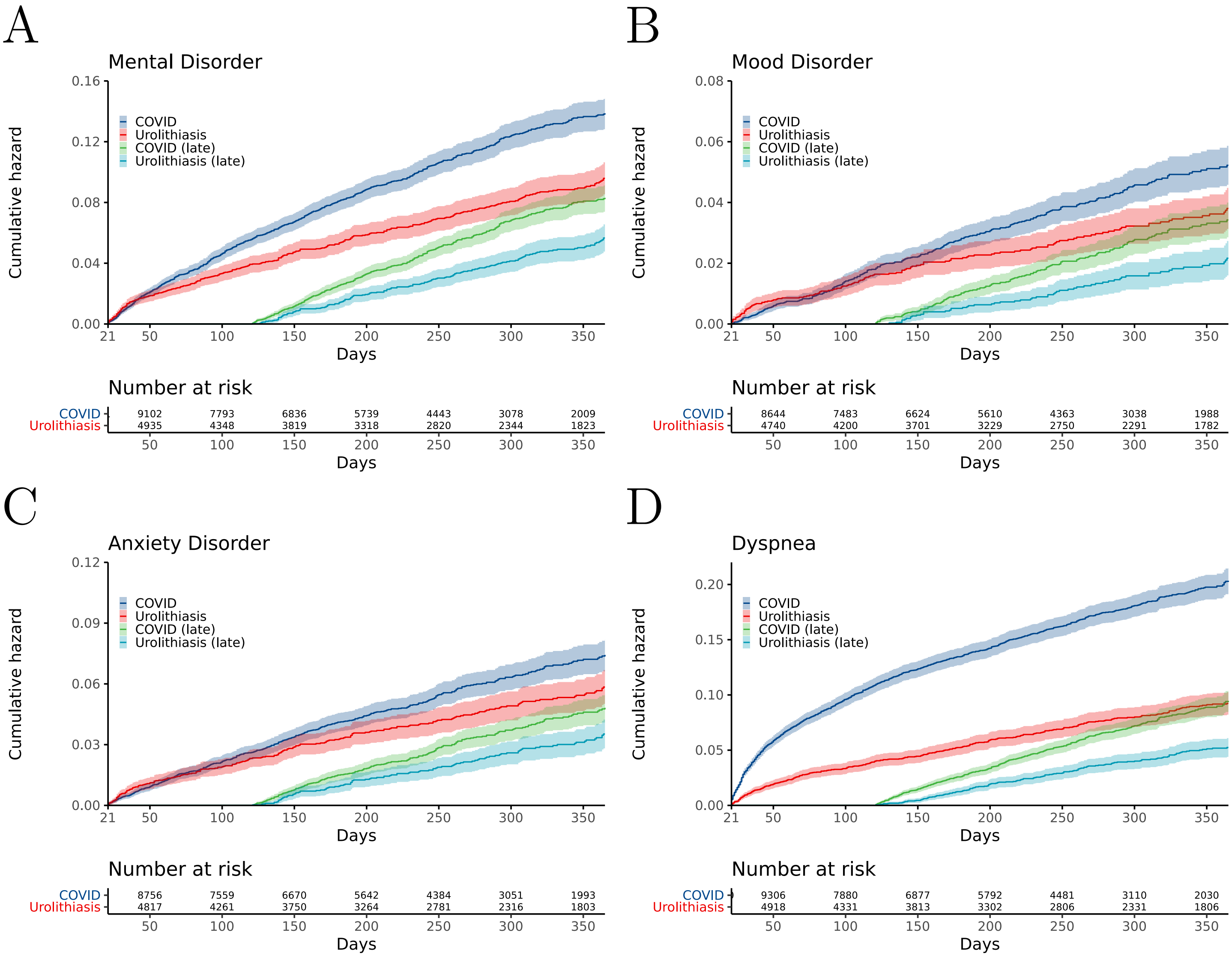


**Figure S3.** Kaplan-Meier curves for onset of first psychiatric diagnoses after COVID-19 diagnosis compared with urolithiasis.

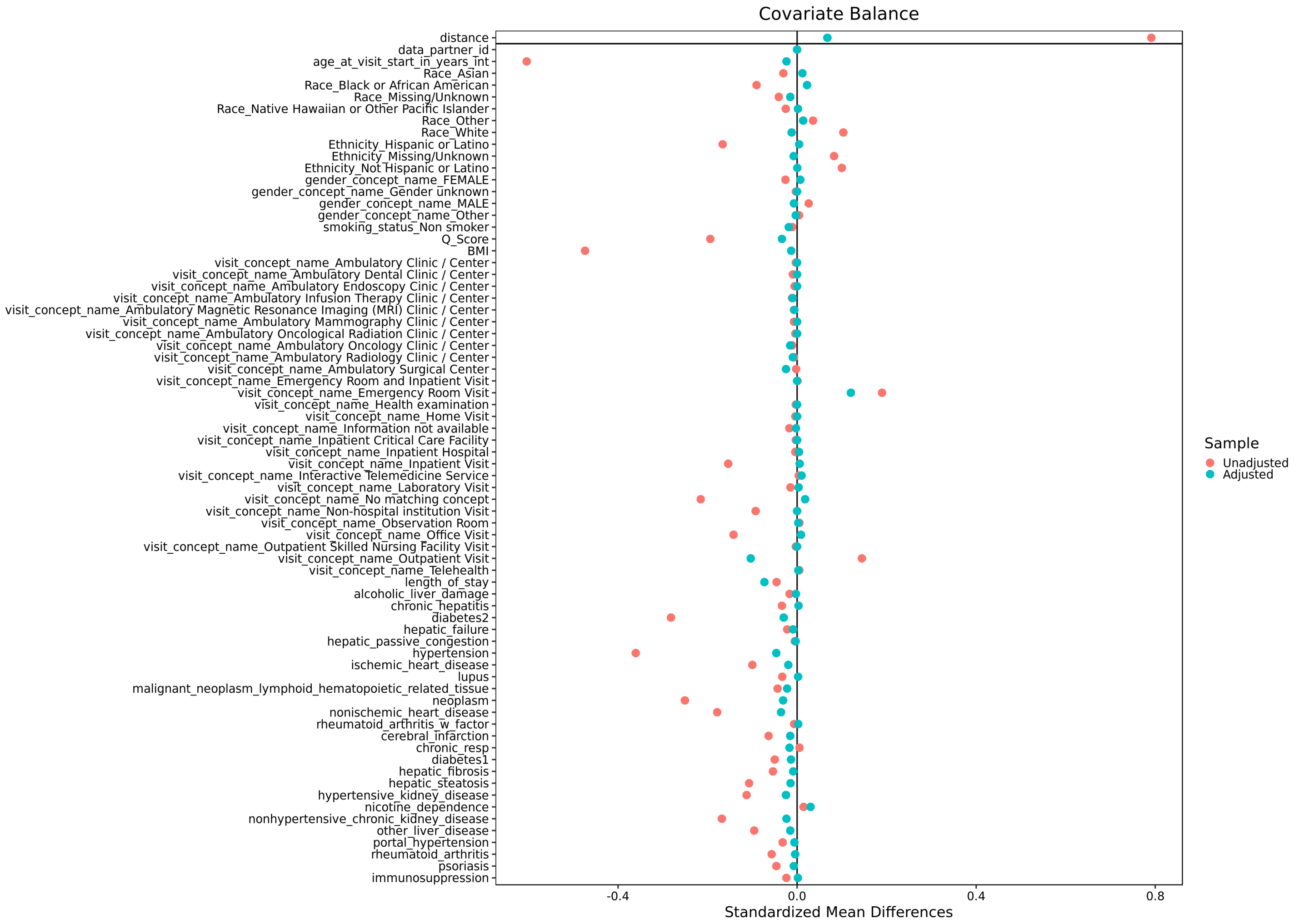


**Figure S4.** Love plot showing the results of propensity score matching for COVID-19 and other respiratory tract infections (RTI). Propensity matching of other groups showed in comparable results.

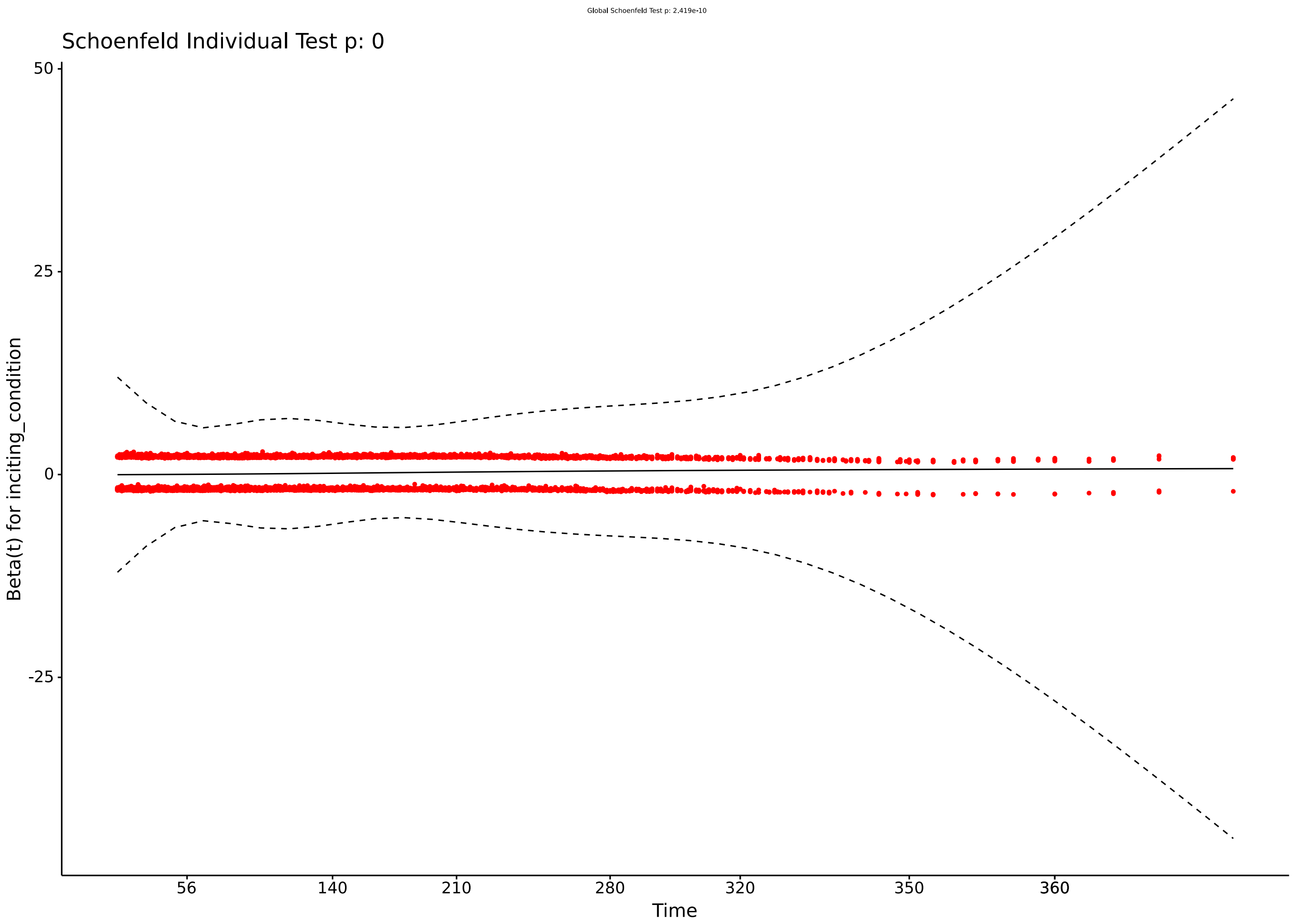


**Figure S5.** Shoenfeld residual plot for COVID-19 and other respiratory tract infections (RTI).

| Outcome | COVID-19 | Fracture | HR | HR $p$-val | PH $p$-val |
|---|---|---|---|---|---|
| | | **120 Days** | | | |
| All psychiatric illness | 5.3% (4.7-5.9) | 3.8% (3.4-4.3) | 1.4 (1.2-1.6) | 0.00040 | 0.19 |
| Mood disorder | 1.6% (1.2-1.9) | 1.3% (1.0-1.6) | 1.2 (0.91-1.7) | 0.17 | 0.09 |
| Anxiety | 2.6% (2.2-3.1) | 1.5% (1.2-1.9) | 1.7 (1.3-2.3) | <0.0001 | 0.70 |
| Fatigue | 6.8% (6.1-7.5) | 3.9% (3.4-4.4) | 1.8 (1.6-2.2) | <0.0001 | 0.0082 |
| Dyspnea | 9.2% (8.4-9.9) | 2.4% (2-2.8) | 4.0 (3.4-4.9) | <0.0001 | 0.026 |
| | | **365 Days** | | | |
| All psychiatric illness | 12.9% (11.7-14.0) | 10.3% (9.3-11.3) | 1.3 (1.0-1.5) | 0.014 | 0.27 |
| Mood disorder | 5.2% (4.3-6.0) | 3.8% (3.2-4.4) | 1.6 (1.2-2.1) | 0.0034 | 0.37 |
| Anxiety | 7.1% (6.2-8.0) | 4.8% (4.1-5.6) | 1.5 (1.1-1.9) | 0.0038 | 0.21 |
| Fatigue | 14.2% (13-15) | 8.9% (8.0-9.8) | 1.8 (1.4-2.2) | <0.0001 | 0.073 |
| Dyspnea | 16.9% (16-18) | 6.7% (5.9-7.5) | 2.1 (1.7-2.5) | <0.0001 | 0.16 |

**Table S1.** Fracture. HR: hazard ratio; PH: proportional hazards

| Outcome | COVID-19 | Urolithiasis | HR | HR $p$-val | PH $p$-val |
|---|---|---|---|---|---|
| | | **120 Days** | | | |
| All psychiatric illness | 5.4% (4.9-5.9) | 3.9% (3.3-4.4) | 1.4 (1.2-1.6) | 0.00017 | 0.0077 |
| Mood disorder | 1.8% (1.5-2.1) | 1.6% (1.3-2) | 1.0 (0.8-1.4) | 0.73 | 0.029 |
| Anxiety | 2.6% (2.2-2.9) | 2.3% (1.9-2.7) | 1.1 (0.88-1.4) | 0.40 | 0.077 |
| Fatigue | 6.6% (6.1-7.1) | 3.7% (3.2-4.2) | 1.8 (1.5-2.1) | <0.0001 | 0.26 |
| Dyspnea | 10.3% (9.7-11) | 3.9% (3.4-4.5) | 2.6 (2.3-3.1) | <0.0001 | 0.10 |
| | | **365 Days** | | | |
| All psychiatric illness | 13% (12.1-13.8) | 9.1% (8.2-10.1) | 1.5 (1.3-1.8) | <0.0001 | 0.15 |
| Mood disorder | 5.1% (4.5-5.7) | 3.7% (3.1-4.4) | 1.7 (1.2-2.2) | 0.0010 | 0.29 |
| Anxiety | 7.2% (6.5-7.9) | 5.7% (4.9-6.5) | 1.4 (1.1-1.8) | 0.0070 | 0.49 |
| Fatigue | 15% (13.8-15.7) | 8.8% (7.9-9.8) | 1.7 (1.4-2.1) | <0.0001 | 0.33 |
| Dyspnea | 18% (17.4-19.3) | 8.8% (7.9-9.7) | 1.7 (1.4-2.1) | <0.0001 | 0.39 |

**Table S2.** Urolithiasis. HR: hazard ratio; PH: proportional hazards

| Predictor | HR (95% CI) w/o freq | HR (95% CI) w freq |
|---|---|---|
| COVID | 1.3 (1.2-1.4) | 1.2 (1.1-1.3) |
| BMI | 1.0 (1.0-1.0) | 1.0 (1.0-1.0) |
| Non-smoker | 0.74 (0.68-0.81) | 0.71 (0.65-0.77) |
| Carleson comorbidity | 1.1 (1.1-1.1) | 1.1 (1.0-1.1) |
| age | 1.0 (1.0-1.0) | 1.0 (1.0-1.0) |
| African American | 1.5 (1.1-2.0) | 1.5 (1.1-2.0) |
| White | 1.7 (1.2-2.3) | 1.6 (1.2-2.2) |
| Outpatient | 0.62 (0.55-0.70) | 0.64 (0.56-0.72) |
| Not hispanic | 1.2 (1.1-1.4) | 1.2 (1.1-1.4) |
| Male | 0.73 (0.68-0.80) | 0.71 (0.66-0.77) |
| visit frequency | - | 3.9 (3.6-4.2) |

**Table S3.** Predictors from regression with and without post-COVID visit frequency. The major predictors are shown together with their 95% confidence intervals. Cox regression was performed in the same way in both experiments except that visit frequency was included in only one experiment. Visit frequency was defined as the number of visits starting at day 21 following the initial event (diagnosis of COVID-19 or RTI). The number of visits was counted up to the final event in the survival curve (either diagnosis of a mental illness or the last day of observation), and was divided the the total number of days between day 21 and the final event.

### S1 Supplementary Note: Code documentation

This project was conducted in the National Institute of Health (NIH) N3C Data Enclave on the Palantir Foundry platform (Palantir Technologies Inc., Denver, Colorado). This platform organizes code in nodes (transformations) that form a directed acyclic graph and cannot easily be presented in linearized fashion. The purpose of this documentation is to illustrate the approach taken by our analysis and to promote reproducibility and extensibility on the Palantir platform or (with appropriate changes) on other platforms.

The Palantir platform enables each node to use a different programming language. We chose SQL, Python, and R according to which technique was best adapted to the task at hand.

The N3C Data Enclave (hereafter "Enclave") contains data from over 2.4 million COVID-19 positive patients from 65 health systems in the United States. The dataset utilized in this study was frozen on October 20, 2021. This dataset consists of 7,139,696 patients. Data were harmonized, integrated, and mapped with the Observational Medical Outcomes Partnership (OMOP) 5.3.1 vocabulary [3].

#### S1.1 Data Preparation

The first step of our analysis consisted in the definition of cohorts and the preparation of data needed for the statistical analysis.

#### S1.2 Inputs

The inputs for our analysis included OMOP tables as well as some tables provided by the Palantir platform with processed data.

##### S1.2.1 OMOP Table

The following OMOP tables were used for our analysis (Table S4). We refer the reader to the original OMOP documentation for more information details [4].

| Table | Summary |
|---|---|
| `condition_occurrence` | Dates when a condition is considered to have started and (if applicable) ended |
| `observation_period` | Defines the time period for which a patient's demographics, conditions, procedures and drugs are recorded in the source system with the expectation of a reasonable sensitivity and specificity. |

**Table S4.** OMOP tables used in this study.

In the code workbook, we refer to the `observation_period` table as `Observation_period_oct21` and `condition_occurrence` as `Condition_occurrence_oct21` because we used the October 21, 2021 data freeze.

#### S1.3 Tables offered by the Enclave

The following Enclave-specific tables were used for our analysis.

##### S1.3.1 concept_set_members

This table defines the relations between the codesets and concept sets (Table S5).

In the code workbooks, we refer to this table as `Concept_set_members_oct21`, because the October 21, 2021 data freeze was used.

| codeset_id | concept_id | concept_set_name | concept_name |
|---|---|---|---|
| 837398380 | 436540 | Fracture | Open fracture axis |
| 837398380 | 436824 | Fracture | Open fracture of cervical vertebra without spinal cord injury |
| 837398380 | 436826 | Fracture | Closed fracture of shaft of radius |
| 837398380 | 436832 | Fracture | Closed fracture of sternum |
| 837398380 | 437117 | Fracture | Open fracture of intracapsular section of femur |

**Table S5. concept_set_members table**. Some examples from this table, which is used to coordinate codesets created by N3C Enclave members (often using the OHDSI Atlas tool). The table has three addition columns (not shown here) that specify the version and were used to extract the latest versions for analysis.

##### S1.3.2 complete_patient_table_with_derived_scores

This table is prepared as described [5], and contains derived information that was used to provide information about some of the covariates used in our analysis (Table **S6**).

| Column | data type | comment |
|---|---|---|
| person_id | string | unique patient identifier |
| data_partner_id | integer | identifier of N3C contributing center |
| visit_concept_id | integer | e.g., 9201 (Inpatient Visit) |
| visit_start_date | Date | The date of the initial medical encounter for COVID-19 or control condition |
| visit_concept_name | string | concept name corresponding to visit_concept_id |
| visit_occurrence_id | string | identifier for a visit |
| AKI_in_hospital | string | “Yes” or null |
| ECMO | string | “ECMO” or null |
| Invasive_Ventilation | string | “Invasive Ventilation” or null |
| positive_covid_test | string | “true” or null |
| negative_covid_test | string | “true” or null |
| Suspected_COVID | string | “true” or null |
| in_death_table | string | “true” or null. True if patient known to be deceased |
| age_at_visit_start_in_years_int | integer | age in years |
| length_of_stay | integer | length of stay in days |
| Race | string | e.g., ”Missing/Unknown” |
| Ethnicity | string | e.g., ”Missing/Unknown” |
| gender_concept_name | string | e.g., “FEMALE” |
| smoking_status | string | e.g., “Non smoker” |
| blood_type | string | e.g., “Unknown” |
| covid_status_name | string | e.g., “covid_confirmed_positive” |
| Severity_Type | string | e.g., “Mild” |
| InpatientOrED | boolean | true or false |
| Q_Score | integer | Charlson Comorbidity Index |
| BMI | double | e.g., 18.2 |
| Height | double | in meters |
| Weight | double | in kilograms |
| Testcount | integer | Not used in current analysis |

**Table S6.** Fields of the complete_patient_table_with_derived_scores table

#### S1.4 Data Preparation

The following subsubsections show the code used for preparing the data for analysis. Each subsubsection corresponds to a single node (transform) in the Enclave.

##### S1.4.1 Raw_Comorbidities_Table

**Inputs:**

- `Concept_set_members_oct21` (Section S1.3.1)
- `Condition_occurrence_oct21` (Section S1.2.1)

Here, we first retrieve the covariate `codeset_id`'s from the OMOP `concept_set_members` table (October 21, 2021 data freeze) (lines 1-4). Then we select all condition occurrences corresponding to these concept sets (lines 6-10). Finally, we select as covariates conditions that existed prior to the patient getting COVID-19 (or the control condition). The code reports the covariate as null if it occurred after covid onset to avoid eliminating any `person_id` from list. (lines 12-14).

```
1  WITH CSM AS (SELECT *
2  FROM Concept_set_members_oct21
3  WHERE concept_set_name IN ("alcoholic_liver_damage-bdc","alzheimers_disease-bdc","cerebral_infarction-bdc","chronic_hepatitis-bdc","chronic_resp-bdc","dementia_associated_with_another_disease-bdc","diabetes1-bdc","diabetes2-bdc","hepatic_failure-bdc","hepatic_fibrosis-bdc","hepatic_passive_congestion-bdc","hepatic_steatosis-bdc","hypertension-bdc","hypertensive_kidney_disease-bdc","immunosuppression-bdc","ischemic_heart_disease-bdc","lupus-bdc","malignant_neoplasm_lymphoid_hematopoietic_related_tissue-bdc","neoplasm-bdc","nicotine_dependence-bdc","non-hypertensive_chronic_kidney_disease-bdc","non-ischemic_heart_disease-bdc","other_liver_disease-bdc","portal_hypertension-bdc","psoriasis-bdc","rheumatoid_arthritis-bdc","rheumatoid_arthritis_w_factor-bdc","unspecified_dementia-bdc","vascular_dementia-bdc")
4  AND is_most_recent_version = true),
5
6  COV_OCC AS (SELECT person_id, concept_set_name, MIN(condition_start_date) AS start_date
7  FROM Condition_occurrence_oct21
8  INNER JOIN CSM
9  ON CSM.concept_id = Condition_occurrence_oct21.condition_concept_id
10 GROUP BY person_id, concept_set_name)
11
12 SELECT person_id, IF(start_date < visit_start_date, COV_OCC.concept_set_name, null) AS covariate
13 FROM COV_OCC
14 RIGHT JOIN Severity_Table_for_COVID_and_Controls USING(person_id)
```

**Listing 1:** Data preparation: retrieve covariates.

##### S1.4.2 Severity_Table_for_COVID_and_Controls

**Inputs:**

- `Concept_set_members_oct21` (Section S1.3.1)
- `Condition_occurrence_oct21` (Section S1.2.1)
- `Complete_patient_table_with_derived_scores_missing_bmi_sites_removed_oct21`(Section S1.3.2)

We first get `concept_IDs` from predefined concept sets for our control groups. We then select data on covariates from the complete patient table (Section S1.3.2). Finally, we add a column called `inciting_condition` that contains either "COVID" or the name of a control condition.

```sql
WITH CCS AS (
  SELECT concept_id , concept_set_name
  FROM Concept_set_members_oct21
  WHERE concept_set_name IN ("RTI",  "Fracture", "Urolithiasis", "Skin Infection")
  AND is_most_recent_version = true),

ST AS (SELECT cpt.person_id,cpt.data_partner_id, visit_start_date,
 cpt.visit_occurrence_id,positive_covid_test,negative_covid_test,
 Suspected_COVID,in_death_table,age_at_visit_start_in_years_int,length_of_stay,
 Race,Ethnicity,gender_concept_name,smoking_status,covid_status_name,Q_Score,
 BMI,Height,Weight,Testcount,condition_start_date,condition_concept_id, Severity_Type,
 visit_concept_name
FROM Complete_patient_table_with_derived_scores_missing_bmi_sites_removed_oct21 AS cpt
LEFT JOIN Condition_occurrence_oct21 USING(visit_occurrence_id))

SELECT DISTINCT visit_occurrence_id, person_id, IF(positive_covid_test = true, "COVID",
    CCS.concept_set_name) as inciting_condition, data_partner_id,   visit_start_date,
    age_at_visit_start_in_years_int, length_of_stay, Race, Ethnicity, gender_concept_name,
     smoking_status, covid_status_name, Q_Score,BMI,Height,Weight, visit_concept_name
FROM ST
LEFT JOIN CCS
ON CCS.concept_id = ST.condition_concept_id
WHERE ((ISNULL(ST.Suspected_COVID) AND NOT ISNULL(CCS.concept_id))
  OR ST.positive_covid_test = true) AND ISNULL(in_death_table)
  AND Severity_Type != "Dead_w_COVID"
```

**Listing 2:** The SQL command performs a left join on visit_occurence_id, which is is used in the CPTDS table to denote the visit in which acute COVID-19 or a control condition first occurs.

##### S1.4.3 Psych_Conditions

**Inputs:**

- `Concept_set_members_oct21` (Section S1.3.1)
- `Condition_occurrence_oct21` (Section S1.2.1)

This transform gets patients (`person_id`), concept names for the psychiatric outcome variables and their start dates.

```sql
WITH PCD AS (
  SELECT concept_id , concept_set_name
  FROM Concept_set_members_oct21
  WHERE concept_set_name IN ("Dyspnea", "Mental Disorder",  "Dementia",
    "Anxiety  Disorder", "fatigue", "Mood Disorder", "Psychosis")
  AND is_most_recent_version = true)

SELECT person_id, concept_set_name, condition_start_date
FROM Condition_occurrence_oct21
INNER JOIN PCD
ON Condition_occurrence_oct21.condition_concept_id = PCD.concept_id
```

**Listing 3:** Concept IDs for the control conditions.

##### S1.4.4 pivot_table

**Inputs:**

- `Raw_Comorbidities_Table` (Section S1.4.1)

Create pivot table where each column is a covariate.

```
def pivot_table(filter_by_date):
    df = filter_by_date
    data = df.groupBy("person_id").pivot("covariate").count().drop("null")
    return data
```

**Listing 4:** pivot table to have each covariate as a column.

##### S1.4.5 covariate_list

Here we create a pivot table that makes a column for each of the covariates (which were mentioned in rows in the input table). Note that the suffix `-bdc` was used to keep track of codesets for this project (the Enclave stores codesets from all projects in the same place, and so generally suffices are used for project-specific codesets). We did not include some covariates that are related to psychiatric outcomes such as vascular dementia and Alzheimer's disease, so they are removed here.

```
def covariate_list(pivot_table):
    df = pivot_table
    # Replace counts greater than 0 with 1
    def make_binary(inrow):
        # nulls get converted to "nan" with str(x), necessary because of table conversion
    peculiarities
        outrow = [True if str(x) != "nan" else False for x in inrow]
        return outrow
    df.iloc[:,1:] = df.iloc[:,1:].apply(make_binary)
    df.columns = df.columns.str.replace('-bdc', '')
    df.columns = df.columns.str.replace('-', '')
    df = df.drop(columns=["alzheimers_disease", "dementia_associated_with_another_disease"
    , "unspecified_dementia", "vascular_dementia"])
    return df
```

**Listing 5:** pivot table to have each covariate as a column.

##### S1.4.6 eligible_pt_table

Input:

- `Observation_period_oct21` (Section S1.2.1 )
- `Severity_Table_for_COVID_and_Controls` (Section S1.4.2)

We first extract the period of observation (needed for the time to event analysis) in lines 1-4. We then remove patients with less than one year of history prior to the first medical encounter with COVID-19 or control event (This is done because we want to perform an analysis on new-onset psychiatric disease following COVID-19. Although the previous history of one year does not guarantee that there was no previous encounter, we define the filter as such because of the limited amount of history available in the Enclave) (lines 6-14). Note that there were rare end dates higher than 50,000 days that we interpreted as data problems; affected data points were removed. The next stanza selects patients who developed a psychiatric condition 21 days or later after the initial encounter with COVID1-9 or control condition (lines 15-23).

Finally, the resulting cohort of patients is joined to the covariate list in line 34.

```
  WITH OP AS (
2   SELECT person_id, observation_period_end_date, observation_period_start_date
    FROM Observation_period_oct21
4 ),
```

```
P1YH AS (
  SELECT *, DATEDIFF(observation_period_end_date, visit_start_date) AS obs_after_visit
  FROM Severity_Table_for_COVID_and_Controls
  INNER JOIN OP USING(person_id)
  WHERE ((date_sub(Severity_Table_for_COVID_and_Controls.visit_start_date, 364) > OP.
    observation_period_start_date))
  AND
  DATEDIFF(observation_period_end_date, visit_start_date) < 50000
),

BLP AS (
  SELECT person_id
  FROM P1YH
  INNER JOIN Psych_Conditions AS PC USING(person_id)
  WHERE PC.concept_set_name IN ('Mental Disorder', 'Anxiety Disorder', 'Mood Disorder', '
    Dementia')
  AND
  date_add(P1YH.visit_start_date, 21) > PC.condition_start_date
  GROUP BY person_id
),

RBLP AS (
  SELECT *
  FROM P1YH
  ANTI JOIN BLP USING(person_id)
  WHERE obs_after_visit >= 21
)


SELECT *
FROM RBLP
LEFT JOIN covariate_list USING(person_id)
```

**Listing 6:** eligible patient table.

##### S1.4.7 mental_disorder_info

**Input:**

- `eligible_pt_table` (Section S1.4.6)
- `Psych_Conditions` (Section S1.4.3)

This transform identifies patients who were diagnosed with a new-onset psychiatric condition 21 or more days after the start of the medical encounter for COVID-19 or the control condition.

```
WITH PDD AS (
  SELECT eligible_pt_table.person_id,
     Psych_Conditions.concept_set_name AS mental_illness,
     DATEDIFF(condition_start_date, visit_start_date) as psych_condition_date_diff
  FROM eligible_pt_table
  INNER JOIN Psych_Conditions using(person_id)
)

SELECT person_id, mental_illness, MIN(psych_condition_date_diff) AS
    MIN_psych_condition_date_diff
FROM PDD
WHERE psych_condition_date_diff >= 21
GROUP BY person_id, mental_illness
```

**Listing 7:** Gather patients with a diagnosis of psychiatric illness that appears at least 21 days after COVID-19 or the control condition.

#### S1.5 Create Table 1

**Input:**

- `Eligible_pt_table` (Section S1.4.6 )

The following code was used to gather the information used in Table 1 of the manuscript.

The following transform is written as an R function. In the Enclave, each R or Python transform is written as a function that is executed by the system. The print statement on line 14 causes the table to be printed in another dialog for downstream use.

```
1  library(tableone)
2  
3  table_1_cohort_description <- function(Eligible_pt_table) {
4  
5      cto_nonfactor_vars <- c("data_partner_id", "age_at_visit_start_in_years_int","Race","Ethnicity","gender_concept_name","smoking_status","blood_type","InpatientOrED","Q_Score","BMI","has_condition", "inciting_condition","visit_concept_name", "obs_after_visit", "length_of_stay", "alcoholic_liver_damage", "chronic_hepatitis", "diabetes2", "hepatic_failure", "hepatic_passive_congestion", "hypertension", "ischemic_heart_disease", "lupus", "malignant_neoplasm_lymphoid_hematopoietic_related_tissue", "neoplasm", "nonischemic_heart_disease", "rheumatoid_arthritis_w_factor", "vascular_dementia", "alzheimers_disease", "cerebral_infarction", "chronic_resp", "dementia_associated_with_another_disease", "diabetes1", "hepatic_fibrosis", "hepatic_steatosis", "hypertensive_kidney_disease", "nicotine_dependence", "nonhypertensive_chronic_kidney_disease", "other_liver_disease", "portal_hypertension", "rheumatoid_arthritis", "unspecified_dementia","psoriasis", "immunosuppression")
6      cto_factor_vars <- c("Race", "Ethnicity", "gender_concept_name", "smoking_status", "blood_type", "InpatientOrED", "visit_concept_name", "inciting_condition")
7  
8      print(paste(date(), "cohort description"))
9      tab1 <- CreateTableOne(
10          vars = c(cto_nonfactor_vars, cto_factor_vars),
11          data = Eligible_pt_table,
12          factorVars = cto_factor_vars,
13     )
14     print(tab1, showAllLevels = TRUE)
15     return(NULL)
16 }
```

**Listing 8:** Creating table 1 with the tableone package.

#### S1.6 Statistical analysis

The statistical analysis is performed using different transforms for different control groups. Here we show a typical example.

#### S1.7 Create case propensity-matched control cohorts

In the following code, we create case-control datasets for analysis. In the example we show here, we use "RTI" (non-COVID-19 respiratory tract infection); an analogous analysis was performed for the other cohorts described in the main manuscript.

##### S1.7.1 RTI_COVID_Matched

**Input:**

- `Eligible_pt_table` (Section S1.4.6 )

For each of the case control groups, a function called `run_matchit` is called that uses the R package `matchit` to perform propensity score matching [6]. For instance, for the RTI group, the following transform is run.

```
RTI_COVID_Matched <- function(Eligible_pt_table) {
    return(run_matchit(Eligible_pt_table, "RTI", matching_ratio=1))
}
```

**Listing 9:** COVID-19 vs. RTI matched cohort.

A similar analysis was performed for COVID-19 vs. Fracture.

```
Fracture_COVID_Matched <- function(Eligible_pt_table) {
    return(run_matchit(Eligible_pt_table, "Fracture", matching_ratio=1))
}
```

**Listing 10:** COVID-19 vs. Fracture matched cohort.

A similar analysis was performed for COVID-19 vs. Urolithiasis.

```
Urolithiasis_COVID_Matched <- function(Eligible_pt_table) {
    return(run_matchit(Eligible_pt_table, "Urolithiasis", matching_ratio=2))
}
```

**Listing 11:** COVID-19 vs. Urolithiasis matched cohort.

We do not show further code related to Fracture or Urolithiasis because the code is completely analogous to the code for RTI.

##### S1.7.2 run_matchit

We run propensity matching by considering the cohort assignment (COVID-19 vs. RTI) as the "treatment" (i.e., `has_condition`) in preparation for the investigation of whether there is a significant difference between the two groups with respect to time to event of a psychiatric diagnosis.

```
run_matchit <- function(data, inciting_condition_name, matching_ratio=1) {
    set.seed(42)
    drop_inc_rows <- TRUE

    data <- data[(data$inciting_condition == "COVID" | data$inciting_condition == inciting
    _condition_name),]
    factor_cols <- c("Race", "Ethnicity", "gender_concept_name", "smoking_status", "visit_
    concept_name")

    cols_of_interest <- c("data_partner_id", "age_at_visit_start_in_years_int","Race",
    "Ethnicity","gender_concept_name","smoking_status","Q_Score","BMI","has_condition",
    "person_id", "inciting_condition","visit_concept_name", "obs_after_visit",
    "length_of_stay", "alcoholic_liver_damage", "chronic_hepatitis", "diabetes2",
    "hepatic_failure", "hepatic_passive_congestion", "hypertension",
    "ischemic_heart_disease", "lupus",
    "malignant_neoplasm_lymphoid_hematopoietic_related_tissue", "neoplasm",
    "nonischemic_heart_disease", "rheumatoid_arthritis_w_factor",  "cerebral_infarction",
    "chronic_resp",  "diabetes1", "hepatic_fibrosis", "hepatic_steatosis",
    "hypertensive_kidney_disease", "nicotine_dependence",
    "nonhypertensive_chronic_kidney_disease", "other_liver_disease",
    "portal_hypertension",  "rheumatoid_arthritis","psoriasis", "immunosuppression")


    matchit.formula <- as.formula("has_condition ~ data_partner_id +
      age_at_visit_start_in_years_int + Race + Ethnicity + gender_concept_name +
      smoking_status + Q_Score + BMI + visit_concept_name + length_of_stay +
        alcoholic_liver_damage+chronic_hepatitis+diabetes2 + hepatic_failure +
```

```
        hepatic_passive_congestion + hypertension + ischemic_heart_disease + lupus +
         malignant_neoplasm_lymphoid_hematopoietic_related_tissue + neoplasm +
         nonischemic_heart_disease + rheumatoid_arthritis_w_factor +  cerebral_infarction +
         chronic_resp +  diabetes1 + hepatic_fibrosis + hepatic_steatosis +
         hypertensive_kidney_disease + nicotine_dependence +
         nonhypertensive_chronic_kidney_disease + other_liver_disease +
         portal_hypertension + rheumatoid_arthritis  + psoriasis + immunosuppression")

    data$has_condition <- as.integer(data$inciting_condition==inciting_condition_name)

    data <- dplyr::select(data, cols_of_interest)

    for(x in factor_cols){data[[x]] <- as.factor(c(data[[x]]))}

    # assess missingness, drop incomplete rows
    pMiss <- function(x){sum(is.na(x))/length(x)*100}
    print("Missingness:");print(apply(data,2,pMiss))
    print("Missingness pattern:"); print(md.pattern(data))
    if (drop_inc_rows){data <- data %>% drop_na()}

    matched <- MatchIt::matchit(formula = matchit.formula, ratio=matching_ratio,
      method = "nearest", data = data, exact=c( "data_partner_id"),
      caliper=c(age_at_visit_start_in_years_int=5), std.caliper=c(FALSE))
    matched.data <- match.data(matched)

    print(summary(matched))

    m.out <- matched
    print(love.plot(m.out, binary = "std"))

    return(matched.data)
}
```

**Listing 12:** Run matchit.

##### S1.7.3 RTI_matched_w_psych_info

**Input:**

- `RTI_COVID_Matched` (Section S1.7.1)
- `Mental_disorder_info` (Section S1.4.7)

This query merges information from previous transforms and adds a category for the type of visit (inpatient, outpatient, other).

```
SELECT *, IF(visit_concept_name IN ("Inpatient Visit", "Inpatient Hospital",
  "Emergency Room and Inpatient Visit", "Inpatient Critical Care Facility"), "Inpatient",
  IF((visit_concept_name IN ("Outpatient Visit", "Emergency Room Visit","Office Visit",
  "Non-hospital institution Visit", "Observation Room", "Laboratory Visit",
  "Ambulatory Radiology Clinic / Center", "Ambulatory Surgical Center",
  "Ambulatory Infusion Therapy Clinic / Center",
  "Ambulatory Community Health Clinic / Center",
  "Ambulatory Clinic / Center", "Pharmacy visit", "Ambulatory Rehabilitation Visit",
  "Ambulatory Dental Clinic / Center", "Ambulatory Oncology Clinic / Center",
  "Ambulatory Magnetic Resonance Imaging (MRI) Clinic / Center",
  "Ambulatory Mammography Clinic / Center", "Ambulatory Endoscopy Cinic / Center",
  "Mass Immunization Center", "Ambulatory Oncological Radiation Clinic / Center",
  "Outpatient Skilled Nursing Facility Visit", "Health examination")),"Outpatient",
  "Other")) as visit_type
```

```
FROM RTI_COVID_Matched
LEFT JOIN Mental_disorder_info USING(person_id)
```

**Listing 13:** Run matchit.

#### S1.8 coxRegression

**Input:**

- `RTI_matched_w_psych_info` (Section S1.7.3

Here, we perform cox regression to compare time to event (i.e., of a psychiatric diagnosis) of COVID-19 patients vs. a control group. We use short helper functions to avoid code duplication. The following was used for the comparison between COVID-19 and RTI, and analogous drivers were used for the other comparisons.

```
cox_RTI_all_psych <- function(RTI_matched_w_psych_info) {
    return(cox_regression(RTI_matched_w_psych_info, "Mental Disorder", 0.16, "RTI"))
}
```

**Listing 14:** Helper function for Cox regression script.

The code first defines the outcome variable by dividing patients into those who receive a diagnosis of `psych_disorder` as compared to those who did not have any diagnosis (for instance, if we are testing “Mood disorder”, patients with “Anxiety” are excluded from the control group). Then the number of days following the initial medical encounter (COVID-19 or control) is calculated. Cox regression over the entire time course is performed first.

We use the `survSplit` function, which takes a survival data set and a set of specified cut times and splits each record into multiple subrecords at each cut time. This allows us to perform Cox regression separately on this first and the second half of the year following the initial medical encounter. Note that we disregard results after the second cut because of a substantially smaller amount of data. Finally, the code displays a Kaplan Meier plot (this is the code that was used to generate plots shown in the manuscript and supplement).

```
cox_regression <- function(data, psych_disorder, y_limit = 0.08,
    control_condition = "control", cut_point = 120) {
    data$mental_illness[is.na(data$mental_illness)] <- 'none'
    data = data[(data$mental_illness == "none") | (data$mental_illness == psych_disorder)
    ,]
    data$result <- !is.na(data$MIN_psych_condition_date_diff)
    data$time <- pmin(data$MIN_psych_condition_date_diff, data$obs_after_visit, na.rm=TRUE
    )

    # Limit in days for survival analysis
    end_point <- 365


     # Create list of dataframes for second time group
    data_t2 <- data[data$time > cut_point,]
    data_t2$result[data_t2$time > end_point] <- 0
    data_t2$time[data_t2$time >= end_point] <- end_point

    # Get KM fits for two time groups
    km_fit2 <- survfit(Surv(time, result) ~ inciting_condition, data=data_t2)
    km_fit1 <- survfit(Surv(time, result) ~ inciting_condition, data=data)
    km_fit <- list(T1 = km_fit1, T2 = km_fit2)

    # Make temporary plot for risk table for COVID and RTI groups at t1 only
```

```
    temp_plot = ggsurvplot(km_fit1, data=data, title=psych_disorder , xlim=c(21,365),
      risk.table = TRUE, axes.offset=FALSE, conf.int=TRUE, fun='cumhaz', xlab="Days",
      break.x.by=50, font.family = "Arial", fontsize = 5.5,
      ggtheme = theme_classic2(base_size=26, base_family = "Arial"), palette = c("lancet"),
      legend.labs =c("COVID", control_condition), legend.title="")
    my_plot = ggsurvplot_combine(km_fit, data=data, title=psych_disorder , xlim=c(21,365),
      ylim=c(0,y_limit), risk.table = TRUE, axes.offset=FALSE, conf.int=TRUE, censor.shape="",
      fun='cumhaz', xlab="Days", break.x.by=50,
      legend.labs= c('COVID', control_condition, 'COVID (late)', paste0(control_condition, " (late)")),
      ggtheme = theme_classic2(base_size=26, base_family = "Arial"), font.family = "Arial",
      palette = c("lancet"), legend.title="")
    my_plot$plot = my_plot$plot + scale_x_continuous(breaks = c(21,50,100,150,200,250,300,350),
      expand=c(0,0), limits=c(0,365)) + theme(legend.position=c(0.15,0.8))
    my_plot$table <- temp_plot$table

    # Factorize data
    factor_cols = c("Race", "Ethnicity", "gender_concept_name", "smoking_status", "inciting_condition", "visit_type")
      for(x in factor_cols){data[[x]] <- as.factor(data[[x]])}
      cox_fit <- coxph(Surv(time, result) ~ BMI + smoking_status + Q_Score + age_at_visit_start_in_years_int + Race + visit_type + Ethnicity + gender_concept_name + inciting_condition, data=data)

    print(paste(date(), "doing zph"))
    temp <- cox.zph(cox_fit) # apply the cox.zph function to the desired model
    print(temp) # display the results
    print(summary(cox_fit))

    # image: svg
    print(my_plot)

    return(data)
}
```

**Listing 15:** Cox Regression.

#### S1.9 make_table_2_RTI

**Input:**

- `cox_regression` (Section S1.8)
- Analogous inputs for the other comparisons.

The following code combines all of our input datasets and extracts information that is used to generate table 2.

```
make_table_2_RTI <- function(cox_RTI_all_psych, cox_RTI_Mood, cox_RTI_Anxiety,
    cox_RTI_Fatigue, cox_RTI_Dyspnea) {
    sequelae_data <- list(cox_RTI_all_psych, cox_RTI_Mood, cox_RTI_Anxiety,
      cox_RTI_Fatigue, cox_RTI_Dyspnea)
    return(make_table_2(sequelae_data, "RTI", cut_point = 120))
}
```

**Listing 16:** Make Table 2 help function.

The `make_table_2` function is as follows.

```
make_table_2 <- function(sequelae_data, comparator, cut_point) {
    # Set global constants
    end_point <- 365
    round_dec_places <- 1
    condition <- "COVID-19"
    hazard_ratio_name <- "Hazard_Ratio"
    ph_pval<- "Proportional_hazard_p_value"
    hr_pval <- "Hazard_Ratio_P_Value"
    sequelae_names <- c("All psychiatric illness (t1)", "Mood disorder (t1)", "Anxiety (t1)", "Fatigue (t1)", "Dyspnea (t1)", "All psychiatric illness (t2)", "Mood disorder (t2)", "Anxiety (t2)", "Fatigue (t2)", "Dyspnea (t2)")
    # Get hazards at cut_point and end_point
    hazard <- lapply(sequelae_data, function(dat){
        # Run Kaplan-Meier fit
        km_fit <- survfit(Surv(time, result) ~ inciting_condition, data=dat)
        # Get Hazards
        s1 <- summary(km_fit, times = cut_point)
        s2 <- summary(km_fit, times = end_point)
        # Format hazards with confidence interval
        percent1=paste0(round(100*(1-s1$surv), round_dec_places), " (", round(100*(1-s1$upper),round_dec_places), "-" , round(100*(1-s1$lower), round_dec_places), ")")
        km_fit2 <- survfit(Surv(time, result) ~ inciting_condition, data=dat)
        percent2=paste0(round(100*(1-s2$surv), round_dec_places), " (", round(100*(1-s2$upper),round_dec_places), "-" , round(100*(1-s2$lower), round_dec_places), ")")
        # Throw data in a sloppy dataframe
        return.df <- data.frame(
            percent1[1],
            percent1[2],
            percent2[1],
            percent2[2]
        )
        colnames(return.df) <- c(condition, comparator, "cond_t2", "comp_t2")
        return(return.df)
    })
    # Make a nice dataframe
    hazard_df_raw <- do.call(rbind, hazard)
    hazard_df <- hazard_df_raw[,c(condition, comparator)]
    hazard_temp <- hazard_df_raw[,c("cond_t2", "comp_t2")]
    names(hazard_temp) <- names(hazard_df)
    hazard_df <- rbind(hazard_df, hazard_temp)
    # Create list of dataframes for first time group
    sequelae_t1 <- lapply(sequelae_data, function(data) {
        data_t1 <- data
        data_t1$result[data_t1$time > cut_point] <- 0
        data_t1$time[data_t1$time > cut_point] <- cut_point
        return(data_t1)
    })
    # Create list of dataframes for second time group
    sequelae_t2 <- lapply(sequelae_data, function(data) {
        data_t2 <- data[data$time > cut_point,]
        data_t2$result[data_t2$time > end_point] <- 0
        data_t2$time[data_t2$time > end_point] <- end_point
        return(data_t2)
    })
    # Put time groups into single iterable dataframe
    sequelae_tgroups = c(sequelae_t1, sequelae_t2)
    # Test proportional hazards assumption and do Wald test for each sequela and time group
    models <- lapply(sequelae_tgroups, function(dat){
```

```
        # run cox model and get wald test results
        dat$inciting_condition = factor(dat$inciting_condition, levels = c(comparator, "COVID"))
        cox_fit <- coxph(Surv(time, result) ~ inciting_condition + BMI + smoking_status + Q_Score + age_at_visit_start_in_years_int + Race + visit_type + Ethnicity + gender_concept_name , data=dat)
        cox_conf <- summary(cox_fit)$conf.int
        HR <- format(cox_conf["inciting_conditionCOVID", "exp(coef)"], digits=2)
        low_conf <- format(cox_conf["inciting_conditionCOVID", "lower .95"], digits=2)
        high_conf <- format(cox_conf["inciting_conditionCOVID", "upper .95"], digits=2)
        hazard_ratio <- paste0(HR, " (", low_conf, "-", high_conf, ")")
        hazard_ratio_p_value <- summary(cox_fit)$coefficients["inciting_conditionCOVID","Pr(>|z|)"]
        # Run proportional hazard assumption test
        cox_temp <- coxph(Surv(time, result) ~ inciting_condition , data=dat)
        cox_zph <- cox.zph(cox_temp)
        prop_haz_p <- cox_zph$table["inciting_condition","p"]
        # Return dataframe blanks for columns that will be filled with hazards
        return.df <- data.frame(
            "0",
            "0",
            hazard_ratio,
            hazard_ratio_p_value,
            prop_haz_p
        )
        colnames(return.df) <- c(condition, comparator, hazard_ratio_name, hr_pval, ph_pval)
        return(return.df)
    })
    # Create final output dataframe
    df <- do.call(rbind, models)
    df[,condition] <- hazard_df[,condition]
    df[,comparator] <- hazard_df[,comparator]
    row.names(df) <- sequelae_names
    print(df)
    return(df)
}
```

**Listing 17:** Make Table 2.
